# Deep Learning-Driven Pattern Analysis of Dried *E. coli*-Laden Urine Deposits for Point-of-Care Diagnostics

**DOI:** 10.1101/2025.10.14.25337322

**Authors:** M Ashwin Ganesh, M Sophia, Jason Joy Poopady, Abdur Rasheed, Durbar Roy, Amey Nitin Agharkar, Visakh Vaikuntanathan, Kirti Parmar, Dipshikha Chakravortty, Saptarshi Basu

**Affiliations:** Department of Mechanical Engineering, Indian Institute of Science, Bengaluru, Karnataka, India; Aix Marseille University, CNRS, IUSTI, Marseille, France; International Center for Theoretical Sciences, Tata Institute of Fundamental Research, Bengaluru, Karnataka, India; Department of Mechanical Engineering, Shiv Nadar Institution of Eminence, Greater Noida, India; Division of Biological Sciences, Department of Microbiology and Cell Biology, Indian Institute of Science, Bengaluru, Karnataka, India; Adjunct Faculty, School of Biology, Indian Institute of Science Education and Research, Thiruvananthapuram, Kerala, India

**Keywords:** Deep Learning, Morphological Pattern Analysis, Cluster Validity Indices, Point-of-Care Diagnostics, Cyber-Physical Systems.

## Abstract

Urinary tract infections (UTIs) are a rising global health concern, primarily caused by *Escherichia coli (E. coli),* disproportionately affecting women and the elderly. Despite advancements in diagnostic techniques, critical gaps persist: time delays, high costs, and false-positive predictions. This necessitates the development of rapid, reliable, and low-cost point-of-care diagnostic tools. As a vital step towards addressing this need, we present a machine learning-based morphological pattern analysis framework that evaluates dried deposits formed by *E.* co/i-laden urine droplets. Following controlled evaporation, urine samples inoculated with *E. coli* at three distinct concentrations are imaged via brightfield microscopy. The computational objective of this study is twofold. First, we perform supervised ternary classification of the deposits based on bacterial concentration, utilizing a lightweight deep convolutional backbone strictly as a spatial feature extractor. Furthermore, to visually analyze feature cluster distributions, the standardized highdimensional feature embeddings are projected using RCA and t-SNE. Second, beyond pattern classification, we define a Severity Factor by applying internal cluster validity indices (CVIs) to these extracted feature embeddings to evaluate morphological pattern deviation. Ultimately, this study establishes a proof-of-concept methodology for analyzing *E.* coZi-laden dried deposits, offering a robust foundation with potential for integration into cyber-physical systems and real-time point-of-care diagnostics in resource-limited settings.

**Graphical Abstract:** 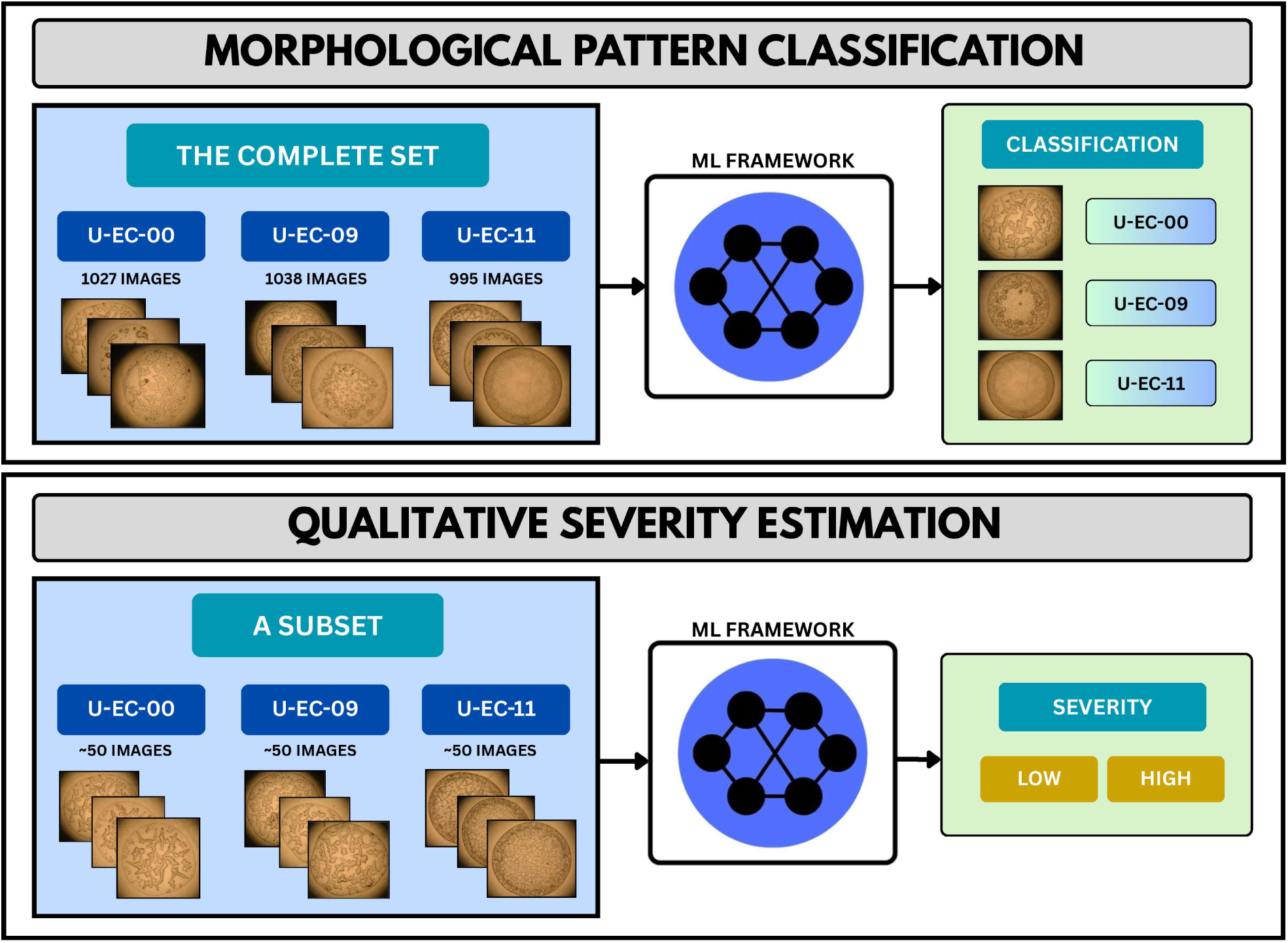

**Highlights:**

- Proposed a morphological pattern analysis framework for dried deposits from *E.* coZz-laden urine droplets.
- Achieved 92.5% accuracy in ternary pattern classification, with MobileNetV2 as the primary feature extractor.
- High-dimensional feature embeddings were projected using PCA and t-SNE to visualize feature clusters.
- Formulated a novel Severity Factor *(S)* using internal Cluster Validity Indices to evaluate morphological deviation.

## 1. Introduction

Urinary Tract Infections (UTIs) are one of the most prevalent bacterial infections worldwide, with over 150 million cases reported annually, predominantly caused by *Escherichia coli (E. coll)* [1]. These infections are significantly more common among adult women, and the risk almost doubles beyond the age of 65 [2]. The most common diagnostic techniques include symptomatic assessment, urinalysis, and traditional urine culture (widely considered the gold standard). However, these traditional methods exhibit notable shortcomings, primarily extended time requirements and high costs [3]. While precise official estimates for UTI prevalence in remote populations remain scarce, multiple global assessments highlight a persistent diagnostic gap at the primary-care level in low-resource settings, driven by factors such as limited laboratory capacity, long turnaround times for urine culture, high out-of-pocket costs, and physical distance to facilities—conditions typical of rural areas that collectively lead to underdiagnosis [4]. Several novel diagnostic approaches, including molecular diagnostics, point-of-care testing (POCT), and artificial intelligence-based techniques, have emerged to overcome these drawbacks [5]. Collectively, these factors highlight the crucial need for developing rapid, low-cost, accessible, and reliable point-of-care diagnostic platforms for UTIs.

Convolutional Neural Network (CNN) and Vision Transformer (ViT)-based architectures are the dominant methods for medical image classification, consistently outperforming traditional machine learning techniques and driving several breakthroughs in the field [6, 7]. ResNet (Residual Neural Network) [8] mitigated the degradation and vanishing gradient problems using skip connections, providing identity shortcuts for gradient flow to prevent information loss. EfficientNet [9] introduced compound scaling (synchronized expansion of a network’s depth, width, and resolution) to overcome the inefficiencies of manual scaling. DenseNet (Dense Convolutional Network) [10] connects each layer to every subsequent layer for enhanced feature propagation and dense feature reuse. MobileNetV2 [11] utilized inverted residuals, depthwise separable convolutions, and linear bottlenecks resulting in reduced computational overhead and memory footprints, making it especially suited for deployment in mobile applications and embedded systems. Vision Transformer (ViT) [12] redefined the state-of-the-art by leveraging global self-attention, a mechanism that captures broad contextual relationships. Following this, several novel architectures such as Swin [13], MaxViT [14], and ConvNeXt [15] were introduced to enhance performance using modernized design principles. Further, hybrid models like Inception-ResNet [16], CoAtNet [17], and MobileViT [18] have demonstrated exceptional capabilities by synergizing the strengths of multiple architectures. However, CNNs were chosen over Transformers as primary feature extractors for this study due to their strong spatial inductive biases. Unlike Transformers, CNNs efficiently preserve fine-grained local features with lower computational complexity, making them particularly effective for robust pattern classification on a limited training dataset [19].

Artificial intelligence—driven medical image analysis has undergone tremendous advancement, with evolving models capable of processing high-volume datasets and recognizing complex, previously undetectable morphological patterns [20]. Within this domain, deep learning architectures have demonstrated exceptional diagnostic capabilities. For example, Zahoor et al. [21] systematically integrated residual and spatial blocks within their Res-BRNet to improve discriminative capacity and generalization for brain tumor classification using Magnetic Resonance Imaging (MRI). Similarly, Esteva et al. [22] achieved dermatologist-level classification of skin cancer, demonstrating that an ImageNet-pretrained Google-Inception-v3 architecture trained on a massive clinical dataset could match the accuracy of board-certified specialists. Azizi et al. [23] proposed a self-supervised learning-based training on a vast, unlabeled medical dataset. Apart from classification, medical image segmentation provides the pixel-level spatial resolution required for precise clinical analysis. Foundational architectures like U-Net [24] utilize skip-connections to successfully preserve fine-grained local details, while hybrid models such as TransUNet [25] integrate transformers to capture long-range global contexts. To further satisfy the high-resolution requirements of biomedical imaging, hierarchical architectures like Swin-Unet [26] have been developed to efficiently model both local and global feature representations. Overall, these rapid architectural advancements highlight the important role of artificial intelligence in transforming medical imaging by automating diagnosis and supporting robust clinical decision-making.

Droplet evaporation-based diagnostics operate by analyzing micro-to nanoliter bio-fluid droplets to infer disease-relevant information from their evaporation residues and dried morphologies [27, 28]. Pathological conditions directly influence the composition of complex bio-colloidal fluids, including blood, saliva, tears, and urine, through alterations in the concentrations and interactions of macromolecules, electrolytes, and cellular constituents [29]. Foundational studies and comprehensive reviews in colloid science establish the diagnostic potential of evaporating biological fluids. These works detail how internal fluid motions, specifically Capillary and Marangoni flows, interact with gelation and crystallization to shape the final dried pattern. Furthermore, they define the standard optical measurement techniques, including brightfield imaging, polarized imaging, and profilometry, required to systematically quantify these distinct morphological signatures [30, 31, 32, 33, 34, 35, 36, 37]. Transitioning specifically to pathogen-fluid interactions, Roy et al. [38] elucidated the evaporation physics of bacteria-laden blood droplets, using theoretically validated experiments to demonstrate how bacterial presence modifies internal flow, evaporation dynamics, and final deposition patterns. Pal et al. [39] examined the temporal evaporation dynamics using time-lapse microscopy, providing evidence that evaporation timescale itself carries an additional diagnostic signal. Hegde et al. [40] utilized non-contact vapor modulation to alter the internal fluid dynamics of evaporating respiratory droplets, demonstrating that controlled flow strictly dictates localized bacterial aggregation and the subsequent deposit pattern formation. Urine, being rich in electrolytes, proteins, and metabolites, produces characteristic dried patterns (concentric rings, crystal fans, dendrites, crack networks) whose features correlate with composition and pathology [41], The application of machine learning and pattern recognition techniques to analyze dried deposit formations is a rapidly emerging field [42], Hegde et al. [43] designed a convolution-based architecture as a point-of-care diagnostic tool to classify dried blood droplet images into seven distinct bacterial infections. Demir et al. [44] studied whole blood and urine droplet samples to detect bladder cancer using pretrained ResNet-based architecture. Similarly, Yang et al. [41] performed label-free urinary protein quantification by employing an Inception-ResNet architecture, positioning it as a promising alternative to traditional diagnostic techniques. Building upon these foundational studies, our present work investigates the variation in residual morphological patterns obtained from *E.* coZz-laden sessile urine droplets at discrete bacterial concentrations, employing a deep learning-based pattern recognition framework.

Machine learning for UTI diagnostics has showcased great potential by serving as a rapid diagnostic tool. Several researchers have demonstrated this by feeding multiple types of input data into deep learning models to make predictions [45] and achieve state-of-the-art performance. For predicting bacteriuria in the emergency department, Sheele et al. [46] utilized patient data as feature variables in an extreme gradient boosting (XGBoost) classifier, which predicted the infection with reasonable accuracy. Farashi et al. [47] used demographic, urine test, and blood test data of patients as features in classifiers, including XGBoost, Light Gradient Boosting Machines (LightGBM), and Random Forest for prediction. A desktop application built by Arches et al. [48] used an image of a urine test strip captured by a smartphone as input, utilizing a CNN for extracting features from colored pads into numerical analyte levels. These features were further fed into a Support Vector Machine (SVM) for the final classification. Liou et al. [49] created a publicly available dataset consisting of microscopic images of liquid urine samples from symptomatic patients. A Patch U-Net model was trained to classify and give the count of granular elements, including white blood cells, red blood cells, and bacteria within them, as a precursor for further diagnosis. A large-volume microscopy-based study by Iriya et al. [50] captured the phenotypic features of bacterial motion, distinguishing them from other non-motile particles, wherein a deep neural network was trained based on numerical features summarizing the bacterial movement. Thereby, such contemporary studies collectively signify the strong potential of machine learning in diagnosing urinary tract infection by leveraging diverse data modalities ranging from structured clinical data and test strip images to the direct microscopic analysis of bacteria and their movement.

Severity estimation of urinary tract infections is inherently complex, depending on a multitude of factors ranging from clinical signs to laboratory data. Clayson et al. [51] evaluated a symptom-based severity factor by creating and validating the UTI Symptom Assessment (UTISA) questionnaire, scoring seven prominent features on a 4-point scale based on intensity and bothersomeness. Similarly, Hay et al. [52] developed a pediatric severity measure based on the presence of seven distinct clinical features. From a diagnostic laboratory perspective, De Rosa et al. [53] introduced a probabilistic approach, combining results from standard urinalysis and automated flow cytometry into a single predictive value called the UTI-Risk Score, based on specific urine constituents. Collectively, these studies highlight the multifaceted nature of UTI severity, traditionally quantified through distinct lenses: patient-reported outcomes, clinical risk stratification, and objective biochemical diagnostics. Building upon these clinical foundations, we derive a novel Severity Factor from a strictly morphological perspective, defining it as a function of pattern deviations from a healthy, baseline dried droplet pattern. We propose that bacterial concentration directly correlates with the degree of morphological pattern deviation within dried urine deposits. To mathematically evaluate this deviation, we employ cluster validity indices (CVIs), which are statistical tools [54] that assess the quality of high-dimensional embeddings by comparing intercluster separation and intra-cluster compactness, translating them into a numerical score. In this study, we utilized a robust subset of internal CVIs that operate without prior knowledge of class labels [55]: the Silhouette Score [56], Davies-Bouldin Index [57], Calinski-Harabasz Index [58], and Centroidal Distance.

In this study, we propose a deep learning-driven framework based on morphological pattern analysis of dried *E.* coZi-laden urine deposits for point-of-care diagnostics. Controlled evaporation experiments were conducted using urine samples inoculated with *E. coli* at three distinct concentrations. The resulting dried deposits were imaged using brightfìeld microscopy. Additionally, Scanning electron microscopy (SEM) was performed to gain physical insights into the impact of bacterial presence on the final micro-structural patterns. Our dataset comprised baseline uninoculated urine alongside samples inoculated at 10^9^ CFU/mL and 10^11^ CFU/mL (designated as U-EC-00, U-EC-09, and U-EC-11, respectively). Our computational analysis is structured around a twofold objective, which forms the primary contributions of this work:

1. **Morphological Pattern Classification:** A deep convolutional backbone was utilized to extract robust spatial features for the supervised classification of morphological patterns based on bacterial concentration. Furthermore, these high-dimensional feature embeddings were visualized using linear and non-linear dimensionality reduction techniques.
2. **Qualitative Severity Estimation:** Moving beyond pattern classification, we introduce a novel Severity Factor. This is achieved by applying internal cluster validity indices (CVIs) to the standardized feature vectors extracted from the trained classification pipeline, mathematically evaluating pattern deviation through cluster analysis.

This unified approach combines pattern classification with severity estimation, offering a robust foundation for diagnostic support at the point of care.

## 2. Methodology

### 2.1. Experiments and Data Acquisition

Urine samples were obtained from 13 subjects in total, with appropriate institutional consent. The experimental procedure involved repeated sampling performed over time. Subjects were instructed to collect their first-morning urine sample in two distinct sterile containers. One container was allocated for routine pathological examination (chemical and microscopic analysis). The second container was reserved for droplet evaporation studies, where the raw urine was transferred into microcentrifuge tubes and inoculated with specific bacterial concentrations to simulate varying degrees of bacteriuria. To maintain a consistent 500 *μ*L total volume across all samples, the baseline uninoculated control (U-EC-00) consisted of 500 *μL* of raw urine. The 10^9^ CFU/mL concentration (U-EC-09) was prepared using 495 *μL* of raw urine with 5 *μ*L of bacterial solution, and the 10^11^ CFU/mL concentration (U-EC-11) using 450 *μL* of raw urine with 50 *μL* of bacterial solution.

The evaporation experiments were conducted under controlled environmental conditions, maintaining a Relative Humidity (RH) of 45 ± 2% and an ambient temperature of 28 ± 2 °C. A 1.5 *μL* droplet of the prepared sample was pipetted onto a coverslip, and its evaporation was monitored using top-view optical microscopy. The resulting final patterns were captured using brightfield microscopy at 10 × magnification. Fig. 1 illustrates the typical temporal variation of the normalized volume of the drying sessile urine droplet, demonstrating an increase in drying time with bacterial presence (at 10^11^ CFU/mL) compared to the uninoculated baseline scenario. The figure also presents the pattern evolution imaged during the evaporation process.

**Figure 1:**
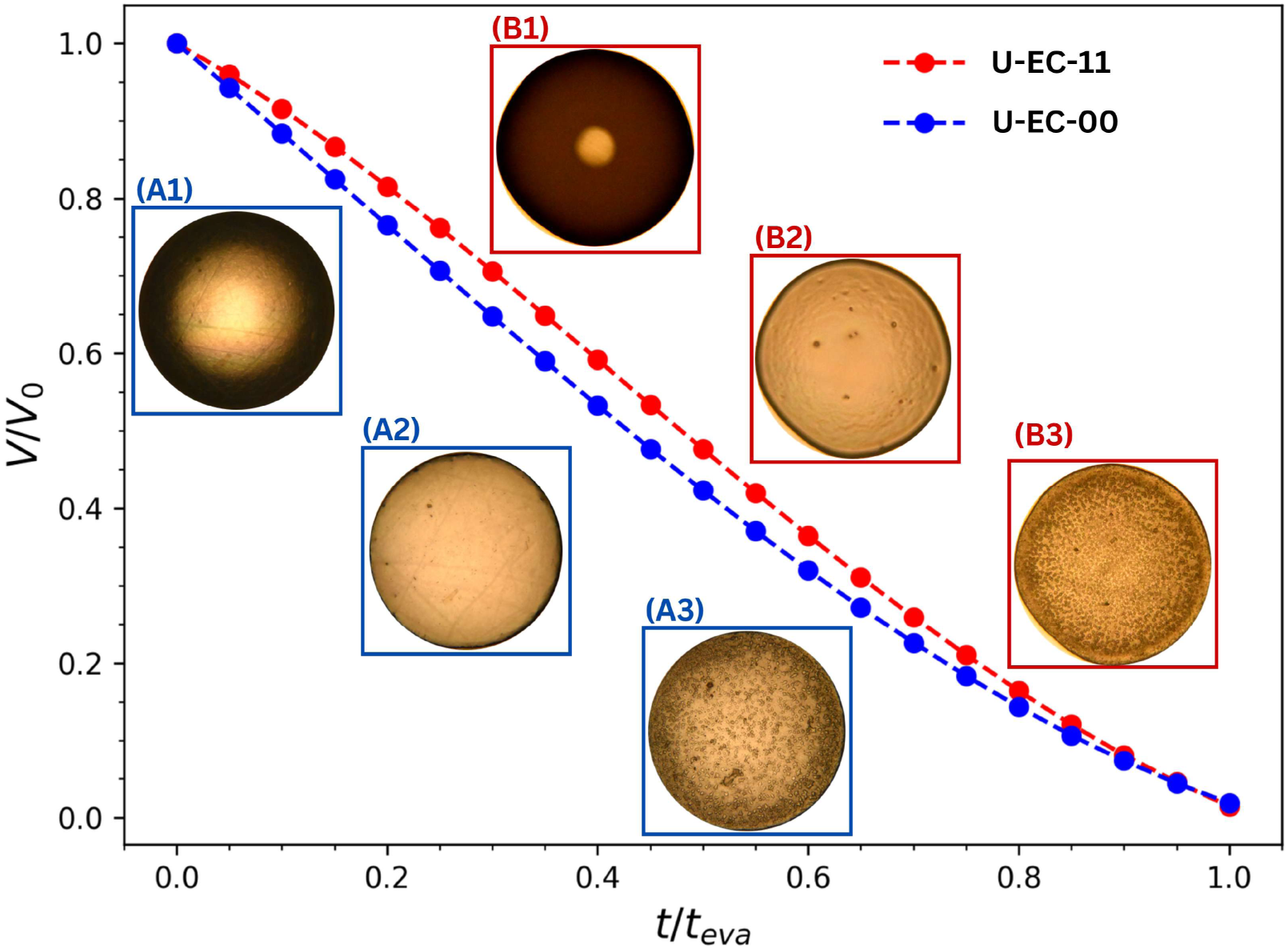
Temporal evolution of the normalized droplet volume during evaporation. Insets illustrate the sequential pattern formation, comparing samples without *E. coli* (panels Al-A3) to samples with *E. coli* at 10^11^ CFU/mL (panels B1-B3).

In addition, scanning electron microscopy (SEM; JEOL-SEM IT 300 Microscopy facility) was employed to characterize the sub-micron architecture of the dried droplet residues at the three *E. coli* concentrations (U-EC-00, U-EC-09, and U-EC-11). Sessile droplets were deposited and dried on standard glass slides (Blue Star, 75 mm × 25 mm). For imaging, the samples were mounted on aluminum stubs using double-sided carbon tape and gold-coated. Backscattered electron (BSE) imaging was conducted in high-vacuum mode with an accelerating voltage of 15 kV, a working distance of approximately 12 mm, and a probe current of 50 nA. Fig. 2 illustrates the morphological details of the patterns, capturing the complete deposit as well as the center and edge sections.

**Figure 2:**
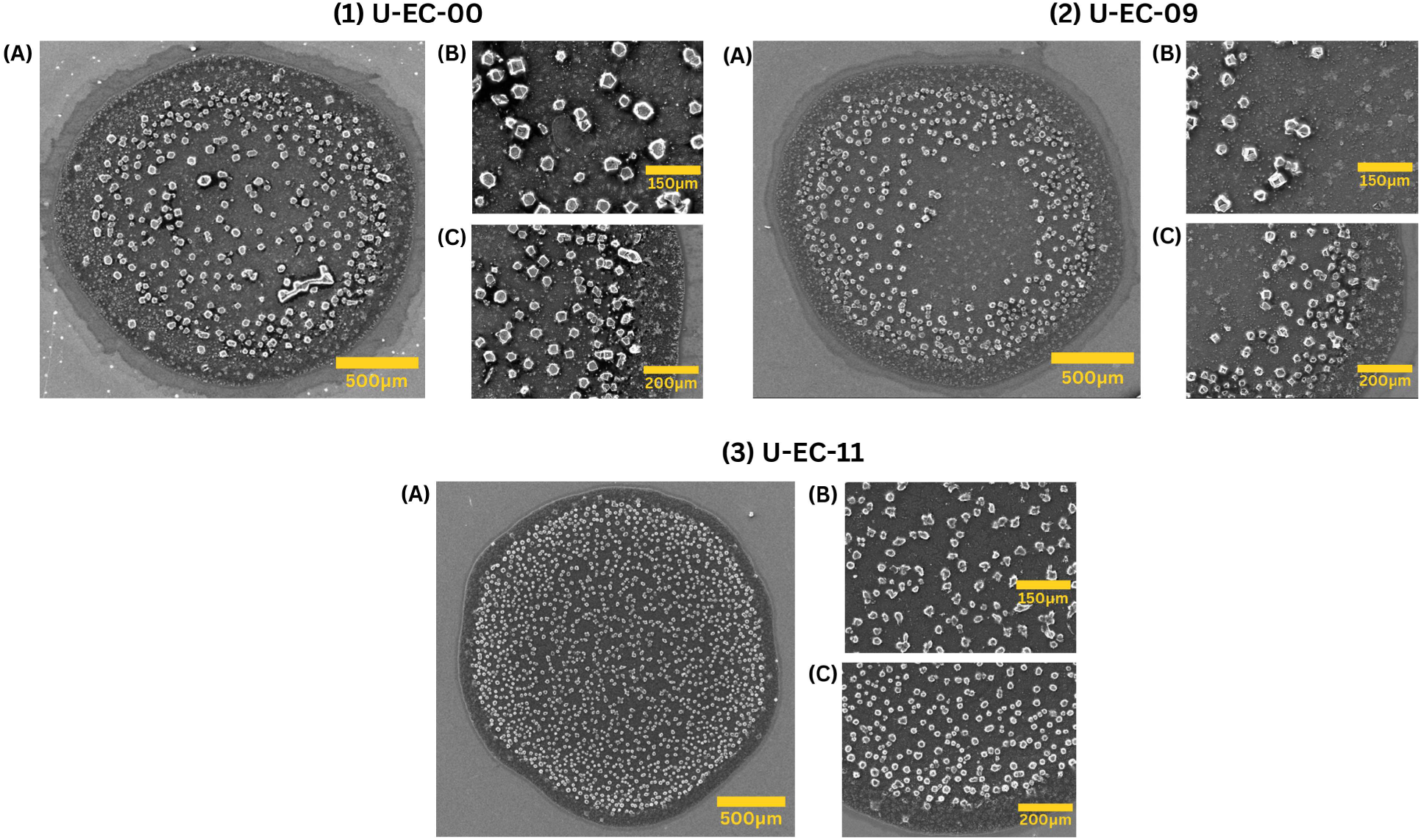
Representative images from Scanning Electron Microscopy of dried patterns formed at various *E. coli* concentrations. Panel (1) shows the pattern without *E. coli* (U-EC-00), panel (2) with *E. coli* at 10^9^ CFU/mL (U-EC-09), and panel (3) with *E. coli* at 10^n^ CFU/mL (U-EC-11). Each panel captures (A) the complete deposit, (B) the central region, and (C) the edge portion of the deposit.

### 2.2. Data Processing

The microscopic images were organized into experimental subsets, where each subset consisted of dried patterns gathered from multiple sessile droplet evaporation runs using the urine sample of a single healthy individual on a specific day. Each subset comprised three distinct bacterial concentration classes: U-EC-00, U-EC-09, and U-EC-11, averaging 50 images per class. This yielded a total of 3,060 images, distributed as 1,027 for U-EC-00, 1,038 for U-EC-09, and 995 for U-EC-11, all stored in high-resolution (6000 × 4000 pixels) RGB format. Our data structuring, along with typical dried patterns observed from three chosen subsets, is depicted in Fig. 3. Crucially, to strictly prevent data leakage, the data were partitioned at the subset level. Of the 18 available subsets, 15 were allocated for training and validation, while the remaining 3 were reserved as a strictly held-out test set. This strategy ensures the test data represent entirely unseen experimental contexts, providing a rigorous benchmark for the system’s robustness.

**Figure 3:**
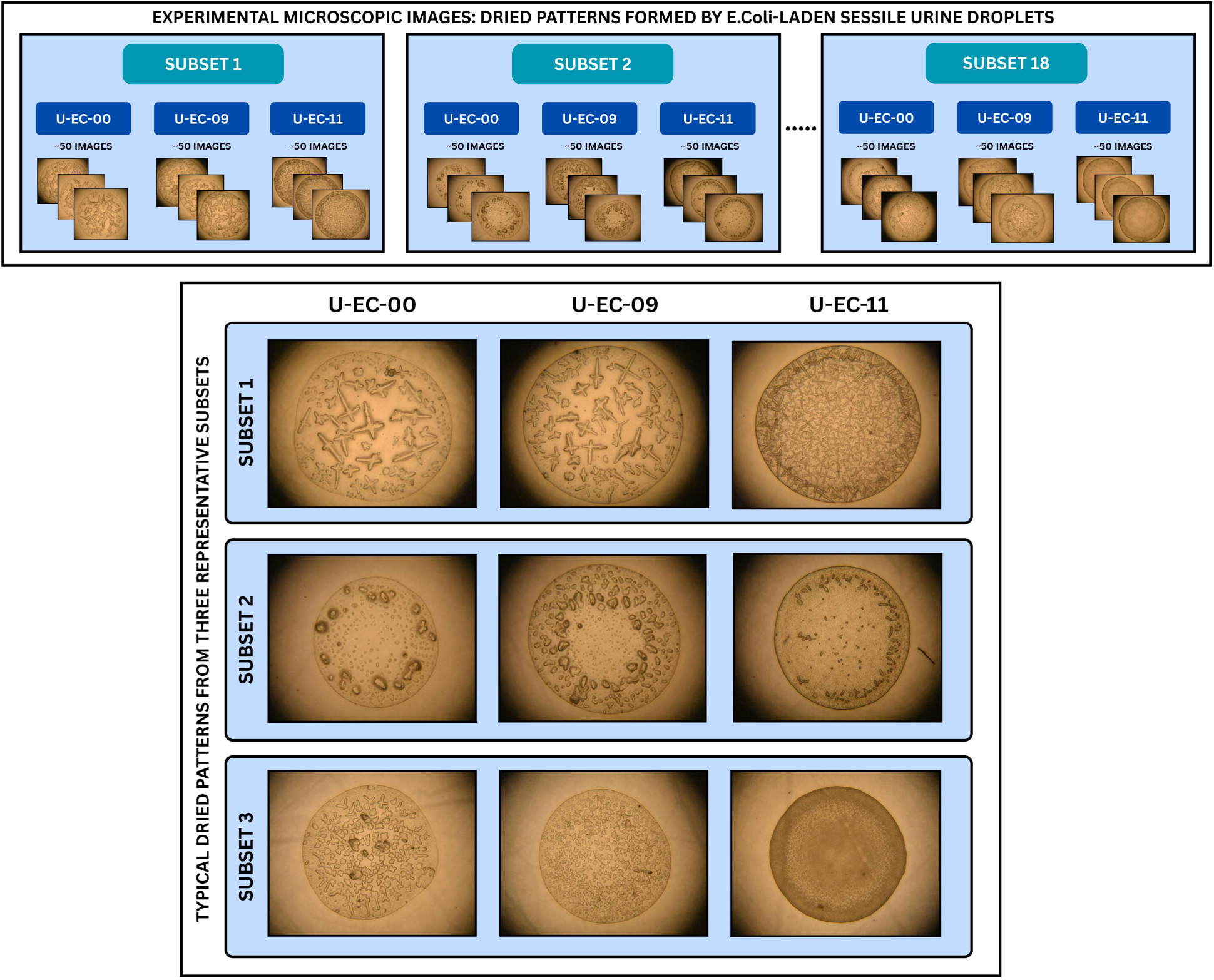
The top panel illustrates a schematic overview of the dataset structure, organized as subsets, each comprising multiple runs at various *E. coli* concentrations. The bottom panel shows representative images of the typical final patterns from three chosen subsets at each bacterial concentration.

Subsequently, an automated image processing pipeline was designed to balance computational efficiency with the preservation of critical spatial features. Within the 15 training subsets, a fivefold stratified cross-validation scheme was implemented to partition the data, ensuring rigorous isolation between training and validation contexts while maintaining consistent class distributions. All images were first subjected to a 2500-pixel center crop to isolate the primary region of interest. They were then resized to a target resolution of 1250 × 1250 pixels using bicubic interpolation with anti-aliasing to preserve fine spatial details without distortion. To mitigate overfitting on our limited dataset, the training folds underwent comprehensive spatial and color augmentations, including random horizontal and vertical flips, rotations up to 180°, and a 10% jitter in brightness, contrast, and saturation. Finally, all images were normalized using standard ImageNet statistics to align with the pretrained convolutional backbone. Specifically, each image pixel *x* was normalized channel-wise as *x’ = (x − μ*)/*σ*, where *μ* = (0.485,0.456,0.406) and *σ* = (0.229,0.224,0.225) for the respective RGB channels. The data were loaded using PyTorch DataLoaders with a batch size of 16 using parallel subprocesses to ensure efficient GPU utilization.

### 2.3. Machine Learning Methodology

Our proposed methodology involves a deep convolutional pipeline designed to achieve our twofold objective: ternary morphological pattern classification and qualitative severity estimation. At a foundational level, CNNs were selected over Vision Transformers (ViTs) as spatial feature extractors because of their inherent inductive biases—specifically, locality and translation invariance. These properties promote robust performance on our limited dataset without necessitating the massive training data typically required by ViTs [12]. Consequently, CNNs reliably extract and preserve the fine-grained spatial features critical for morphological classification. The schematic of this dual-objective architecture is illustrated in Fig. 4.

**Figure 4:**
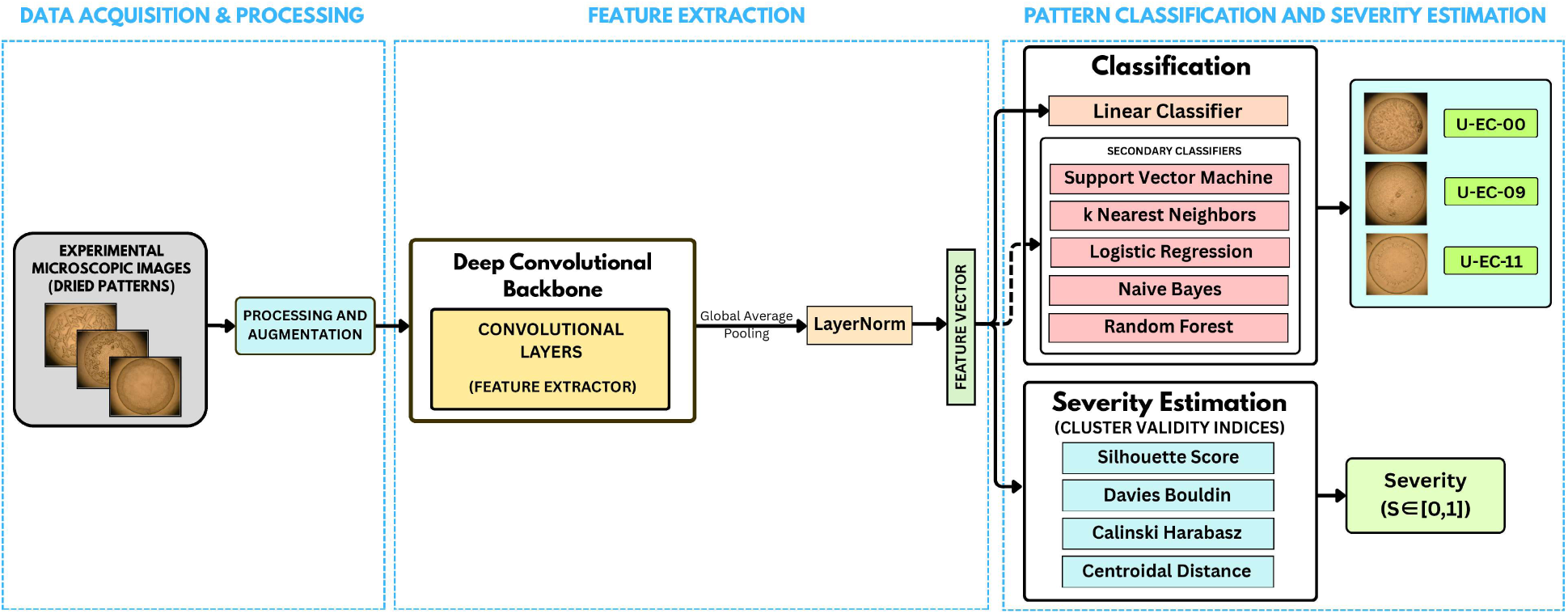
Schematic describing the workflow of our machine learning pipeline for morphological pattern classification and qualitative severity estimation.

Following preprocessing, the images were fed into an ImageNet-pretrained convolutional backbone (with MobileNetV2 acting as the primary architecture) to extract hierarchical spatial features from the dried bacterial deposit images. All deep convolutional backbones were instantiated using the PyTorch Image Models (`timm`) library, utilizing their respective ImageNet-pretrained weights. During the optimization phase, the network was trained end-to-end for 50 epochs, with all convolutional layers in the backbone unfrozen, using the Adam optimizer and categorical cross-entropy loss. The learning rate was regulated by a cosine annealing scheduler, dynamically decaying from 10^-4^ to 10^-6^ to ensure stable convergence. An early stopping mechanism with a patience of 7 epochs was implemented to halt training and retain the model weights corresponding to the minimum validation loss.

Let *X* ∊ ℝ^3×1250×1250^ denote the augmented, preprocessed input image. After spatial feature extraction using a deep convolutional backbone, Global Average Pooling (GAP) was applied, resulting in a 1280-dimensional feature vector:

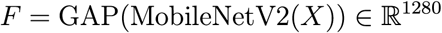

Following this, the extracted feature vector was standardized using Layer Normalization (LayerNorm) to ensure all features share a uniform scale. This step is a mathematical necessity, as it prevents scale-induced bias during dimensionality reduction (PCA/t-SNE), stabilizes the linear and distance-based secondary classifiers, and ensures the accurate computation of Euclidean distances for the CVIs:

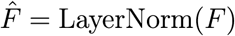

where *F̂* represents the final standardized feature vector utilized for all subsequent assessments.

For pattern classification, the standardized vector *F̂* was passed through a linear classification head incorporating dropout regularization *(p* = 0.5). To rigorously validate the classifier-agnostic robustness of these morphological embeddings, the extracted features were independently evaluated across five diverse algorithms: Support Vector Machine [59], Random Forest [60], Naive Bayes [61], K-Nearest Neighbors [62], and Logistic Regression [63]. To ensure an unbiased comparative baseline, each secondary classifier algorithm underwent exhaustive hyperparameter tuning using a grid search paired with 5-fold cross-validation.

To visually interpret the high-dimensional feature vectors, two dimensionality reduction techniques were employed: linear Principal Component Analysis (PCA) and non-linear t-distributed Stochastic Neighbor Embedding (t-SNE). PCA [64] provides a linear mapping of the feature vectors onto a lower-dimensional subspace defined by the directions of maximum variance. To ensure a consistent representation, this projection was learned strictly from the training data and subsequently applied to the test data. Conversely, t-SNE [65] preserves local data topology by modeling pairwise similarities using Gaussian kernels in the high-dimensional space and a heavy-tailed Student-t distribution in the low-dimensional space. The final visual embedding is computed by minimizing the Kullback-Leibler (KL) divergence between these two distributions over all point pairs.

The predictive capacity of the model on unseen data was evaluated using standard classification metrics, derived from the true positives *(TP_k_),* false positives *(FP_k_*), and false negatives *(FN_k_)* for each class *k.* Precision *(TP_k_/(TP_k_ + FP_k_)* measures the proportion of samples predicted as class *k* that are genuinely correct, while recall *(TP_k_/(TP_k_ + FN_k_))* quantifies the proportion of true class *k* samples correctly identified. The Fl-score combines precision and recall through their harmonic mean to provide a single balanced measure. To assess performance across all classes, we report the macro-average, computed as the unweighted mean of the metrics across all classes. Additionally, the overall accuracy is reported as the proportion of correctly predicted samples across the entire held-out test set.

Moving beyond discrete pattern classification, we derived a qualitative severity factor to mathematically assess the morphological pattern deviation within a given subset. Utilizing the trained classification pipeline, each held-out test subset was processed individually. Images within each subset underwent identical preprocessing and feature extraction to yield standardized feature vectors (F). A cluster analysis was then performed on this feature space to evaluate the morphological pattern overlap across different bacterial concentrations. This was achieved by computing internal CVIs, including the Silhouette Score, Calinski-Harabasz Index, Centroidal Distance, and Davies-Bouldin Index. By mathematically evaluating the degree of feature cluster overlap and distinctiveness, this process yielded a normalized Severity Factor (*S* ∊ [0,1]). The mathematical formulation and significance of the specific CVIs utilized for this analysis are detailed in Table 1.

**Table 1:** Summary of internal cluster validity indices.

| CVI | Definition | Expression | Significance |
| --- | --- | --- | --- |
| **Silhouette Score**<br>(Point Level) | Measures how similar a data point is to its own cluster compared to neighboring clusters, averaged across all points. | $SH = \frac{1}{n} \sum_{i=1}^n \frac{b(i) - a(i)}{\max\{a(i), b(i)\}}$<br>**Range:** $[-1, 1]$ | Near **1**: Dense, well-separated clusters.<br>Near **-1**: Overlapping clusters. |
| **Davies-Bouldin Index**<br>(Cluster Level) | Measures the average similarity of each cluster with its most similar neighbor, based on a ratio of internal spread to external separation. | $DB = \frac{1}{k} \sum_{i=1}^k \max_{j \neq i} \left( \frac{s_i + s_j}{d(c_i, c_j)} \right)$<br>**Range:** $[0, \infty)$ | Near **0**: Superior clustering.<br>**Higher scores**: Poor separation. |
| **Calinski-Harabasz Index**<br>(Global Level) | Measures the ratio of the overall between-cluster variance to the overall within-cluster variance. | $CH = \frac{SS_B / (k - 1)}{SS_W / (n - k)}$<br>**Range:** $[0, \infty)$ | **Higher scores**: Dense, well-separated clusters.<br>**Lower scores**: Indistinct or overlapping clusters. |
| **Centroidal Distance**<br>(Cluster Level) | Euclidean distance between the geometric cluster centroids. | $D_{ij} = d(c_i, c_j)$<br>**Range:** $[0, \infty)$ | **Higher values**: Greater separation between feature clusters. |

**Notations:** $k$ : Number of clusters; $n$ : Number of data points. **Silhouette:** $SH$ : Mean score calculated over all data points $i$ ; $a(i)$ : Mean intra-cluster distance; $b(i)$ : Mean nearest-cluster distance. **Davies-Bouldin Index:** $s_i$ : Average intra-cluster distance. **Calinski-Harabasz Index:** $SS_B$ : Between-cluster sum of squares; $SS_W$ : Within-cluster sum of squares. **Centroidal Distance:** $d(c_i, c_j)$ : Euclidean distance between cluster centroids.

### 2.4. Hypothesis for Qualitative Severity Estimation

To define a quantifiable measure for severity, we introduce a proof-of-concept analysis for computing a severity factor using cluster analysis on the high-dimensional feature embeddings of each independent subset within the unseen test fold. Our approach is predicated on the understanding that urine serves as a highly dynamic biochemical fingerprint [66]. Given that chemical composition fluctuates significantly due to multiple systemic factors (e.g., age, diet, health status, and medication), the resulting dried deposits, and consequently their spatial features, exhibit considerable variability. Based on preliminary visual observations, bacterial presence introduces a distinct morphological deviation on top of the inherent baseline variability. We therefore hypothesize that analyzing this pattern deviation relative to an uninoculated baseline provides qualitative insight into the severity due to the bacterial load. Specifically, we propose that the geometric displacement between *E.* coZi-laden feature clusters and the corresponding uninoculated baseline (U-EC-00) directly correlates with the degree of pattern deviation^1^.

To mathematically evaluate this morphological pattern deviation induced by the bacterial load, internal CVIs were computed on the standardized feature vectors extracted from our deep convolutional pipeline. While a fully unsupervised approach might theoretically provide a more holistic pixel-level model of morphological deviation, explicitly modeling this underlying distribution could result in severe overfitting given the inherent diversity within our limited dataset. Therefore, we utilize the features extracted from our supervised classification pipeline to establish a discriminative model for this morphological deviation, thereby facilitating a quantifiable severity factor. Furthermore, evaluating diagnostic severity based on these features provides a representation of relative categories (e.g., low, medium, or high morphological deviation) rather than an absolute interpretation based on physical magnitudes. Therefore, while the severity factor is mathematically continuous, we formally define it a qualitative metric.

## 3. Results

### 3.1. Insights from Pattern Evolution and SEM Microcharacterization

Fig. 1 contrasts the morphological pattern evolution of evaporating droplets with (panels Bl–B3) and without (panels Al–A3) bacterial presence. The outer ring formation in an evaporating droplet with a pinned contact line is driven by radially outward capillary flow [67]. Dictated by this convective transport mechanism, the bacterial mass is swept toward the contact line, resulting in a significantly thicker ring. Consequently, U-EC-11 exhibited increased peripheral deposition compared to the baseline (U-EC-00) during pattern evolution, forming a structurally denser and more intense edge band (inset B2) [68]. Furthermore, SEM images of the final dried deposits, as presented in Fig. 2, reveal that elevated bacterial concentrations correlate with an increased microcrystal count, indicating enhanced nucleation activity. This microcrystal density enhancement can be attributed to extracellular polymeric substances (EPS) secreted by bacteria, which provide favorable binding sites for crystallizing ions [69, 70]. Crucially, it must be emphasized that these were broadly observed trends from our experiments. Given the high variability of human urine composition [66, 71], these specific insights cannot be universally generalized across all scenarios. This challenge of generalizability in the observed deposits necessitated the adoption of a machine learning-based approach for robust pattern recognition.

### 3.2. Morphological Pattern Classification

The ternary classification of morphological patterns from *E.* cofø-laden sessile urine droplets achieved robust predictive performance using a linear classifier head trained on the standardized feature space extracted by the MobileNetV2 backbone. Evaluated on the held-out test set (Fig. 5A), the model yielded an overall accuracy of about 92.5%, with macro-averaged precision, recall, and Fl-scores of 92.78%. Class-wise evaluation (Table 2) and the row-normalized confusion matrix (Fig. 5B) demonstrated a direct correlation between predictive accuracy and bacterial concentration. The highest concentration class (U-EC-11) attained near-perfect classification, whereas the baseline (U-EC-00) and intermediate (U-EC-09) classes exhibited overlapping feature clusters, resulting in relatively lower, yet robust, discriminatory performance.

**Figure 5:**
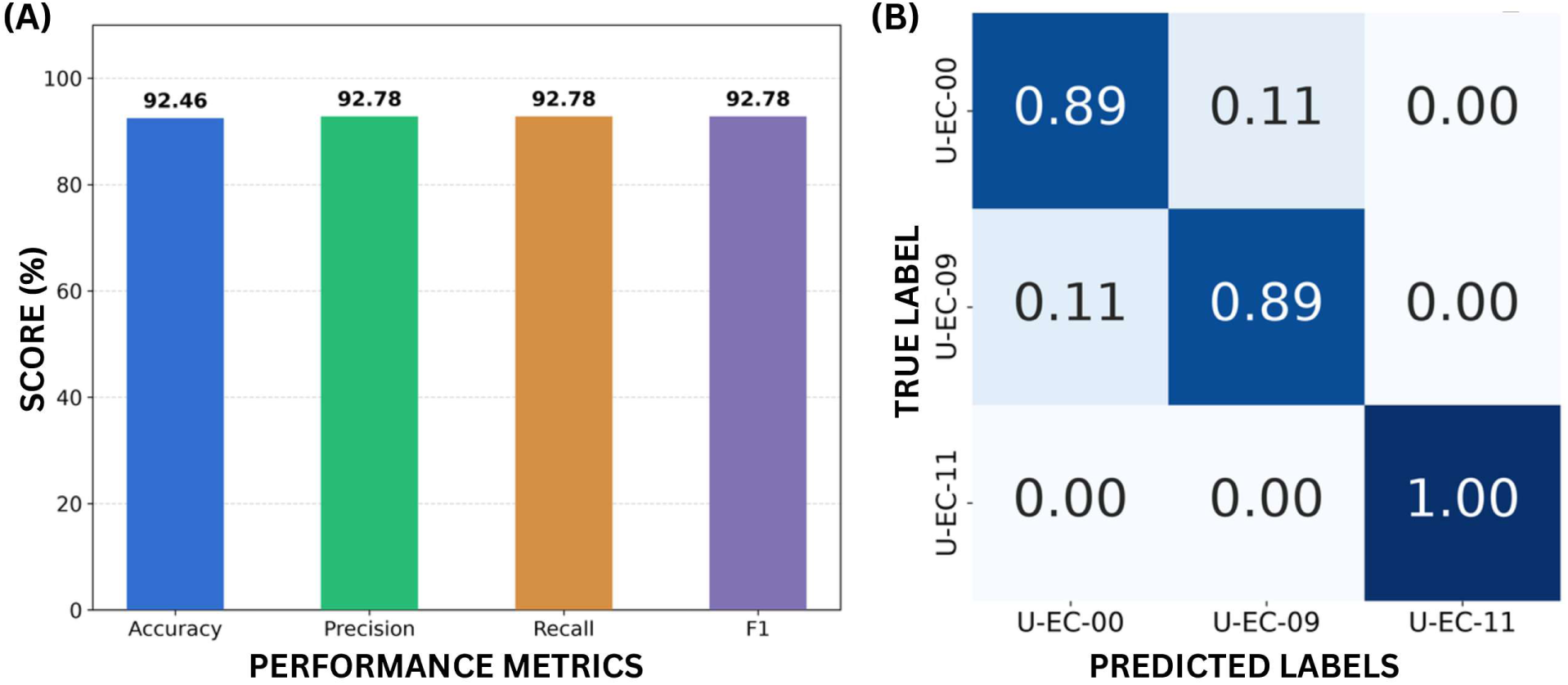
(A) The macro-averaged performance metrics and (B) the row-normalized confusion matrix for morphological pattern classification, based on the linear classifier head.

**Table 2:**
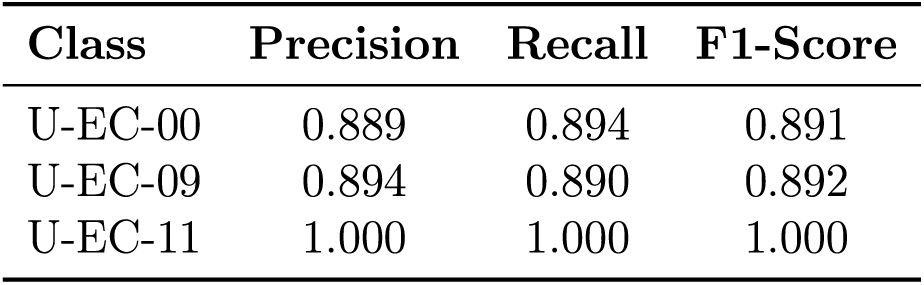
Class-wise performance metrics utilizing the linear classifier head.

To visually interpret the global class separability of the high-dimensional embeddings, two-dimensional PCA and t-SNE maps of the standardized feature vectors are presented in Fig. 6. PCA, which preserves the global variance of the latent space to reveal overall feature clusters at a global level, clearly demonstrated strong linear separability for the highest concentration (U-EC-11). Conversely, U-EC-00 and U-EC-09 formed partially overlapping clusters, accounting for their relatively lower classification metrics. Furthermore, the non-linear t-SNE projection, which probabilistically preserves local neighborhood relationships, revealed fine-grained structures at a subset level. Specifically, distinct sub-clusters emerged within each bacterial concentration, indicating grouping corresponding to each independent test subset, despite the presence of localized regions of inter-class overlap (as discussed in detail in Section 4.1).

**Figure 6:**
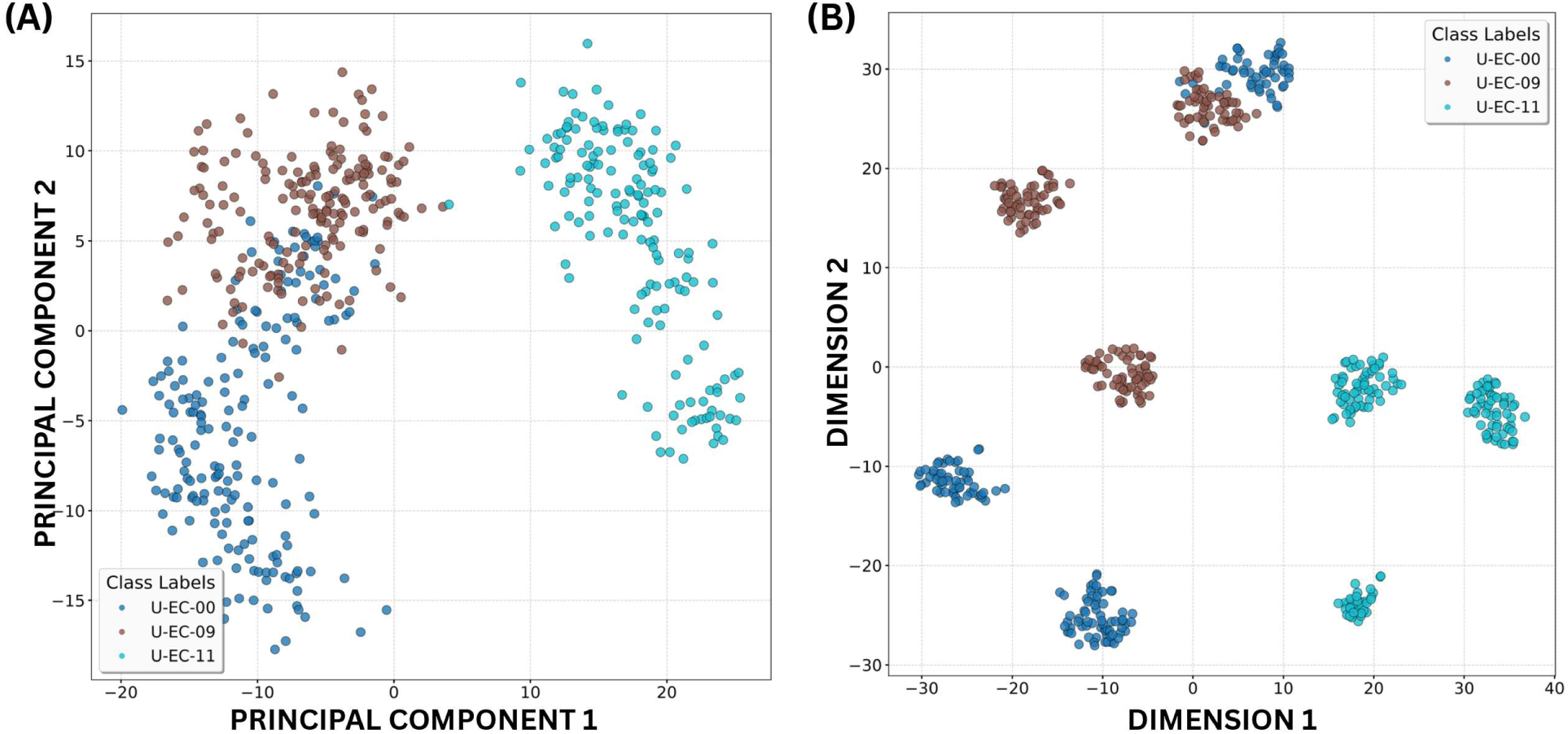
Two dimensional (A) Principal Component Analysis (PCA) and (D) t-SNE plots based on the standardized feature vectors.

To validate the classifier-agnostic robustness of the extracted feature embeddings, the standardized feature vectors were independently evaluated across a suite of secondary machine learning algorithms (Table 3). Predictive performance remained consistently high across all selected algorithms, with Random Forest achieving the highest overall accuracy of 90.3%. Collectively, the efficacy of our morphological pattern classification is comprehensively validated through three complementary approaches: quantitatively through overall and class-wise performance metrics, visually using low-dimensional embeddings of the standardized feature vectors, and comparatively based on multi-classifier performance.

**Table 3:** Comparison of macro-averaged performance metrics evaluated across multiple classifiers using the standardized feature embeddings obtained from the MobileNetV2 model on the held-out test fold.

| Classifier | Accuracy | Precision | Recall | F1-Score |
| --- | --- | --- | --- | --- |
| Support Vector Machine | 0.896 | 0.902 | 0.900 | 0.898 |
| Logistic Regression | 0.863 | 0.900 | 0.865 | 0.867 |
| K-Nearest Neighbors | 0.896 | 0.904 | 0.899 | 0.900 |
| Random Forest | 0.903 | 0.920 | 0.910 | 0.901 |
| Naive Bayes | 0.878 | 0.889 | 0.883 | 0.882 |

### 3.3. Qualitative Severity Estimation

Advancing beyond pattern classification, we introduce a novel severity factor. For each independent subset from the held-out test data, standardized feature vectors *(F̂)* were extracted using the trained deep convolutional pipeline. The degree of morphological pattern deviation was then mathematically evaluated by applying internal CVIs (detailed in Table 1) to these high-dimensional embeddings. Specifically, the overlap in the feature cluster embeddings of the *E.* co/Tladen classes (U-EC-09 and U-EC-11) was computed pairwise against the uninoculated baseline (U-EC-00). Mathematically, higher Silhouette scores, Calinski-Harabasz indices, and inter-cluster Centroidal Distances, coupled with lower Davies-Bouldin indices, signify pronounced morphological deviation and high separability within the standardized feature space. This sub-section details the results for a representative case (Test Subset 1, Fig. 7).

**Figure 7:**
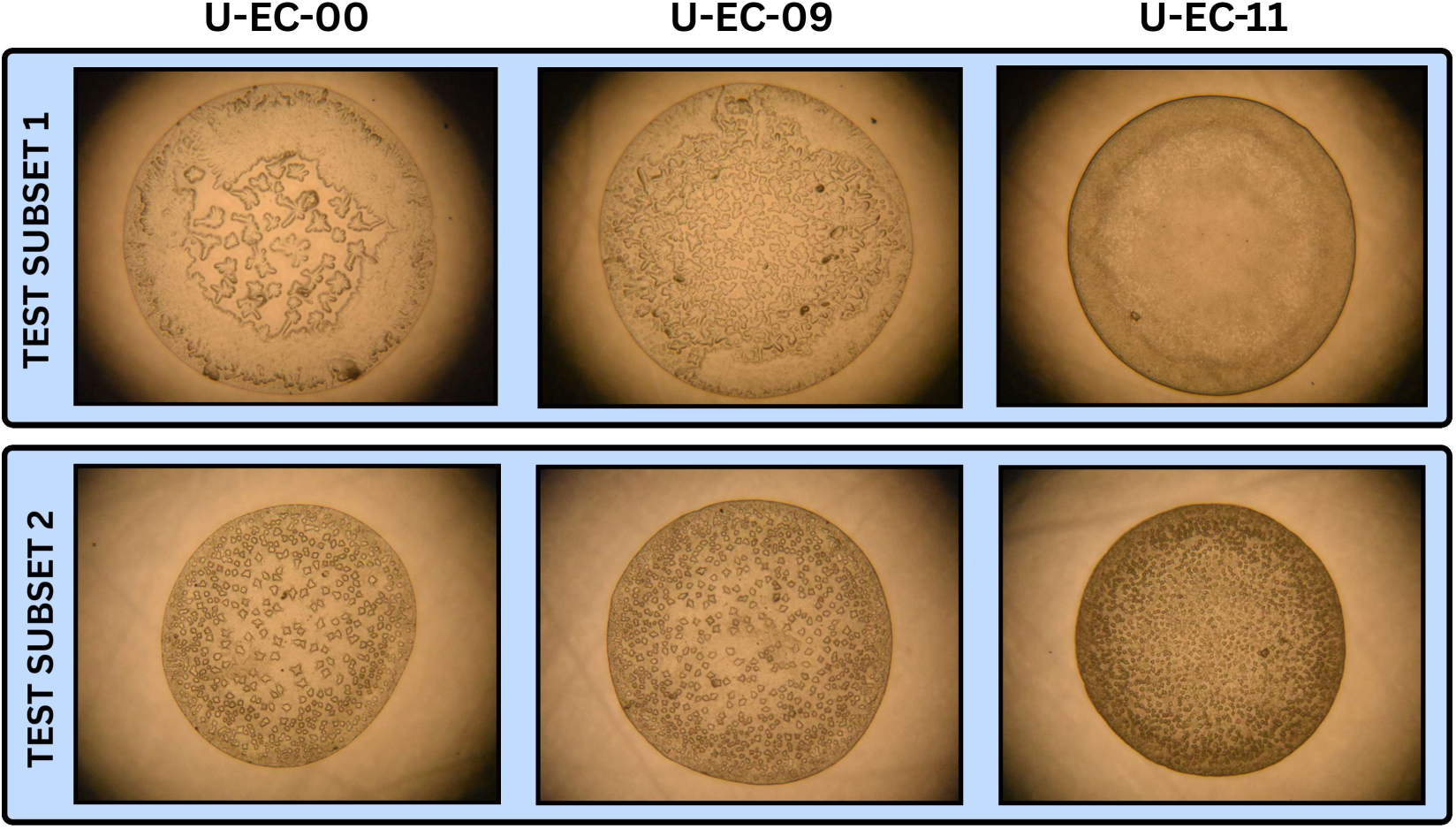
Representative patterns from the two selected test subsets.

Ultimately, the severity factor has been formulated and interpreted according to the hypothesis established in Subsection 2.4. While it is widely acknowledged that no single Cluster Validity Index (CVI) performs optimally across all data distributions [55], the Silhouette score *(SH)* has been explicitly selected for its unique advantages over unbounded global metrics, such as the Calinski-Harabasz and Davies-Bouldin indices. These advantages include its capacity for point-level assessment, its strictly bounded range of [–1,1], and its direct interpretability regarding intra-cluster cohesion and inter-cluster separation. To compute the Severity Factor (S), the Silhouette score *(SH)* was subjected to min-max normalization, mapping it to a strictly positive range of [0,1]. Consequently, an *S* value approaching 0 indicates significant overlap (minimal morphological deviation from the baseline), whereas an *S* value approaching 1 denotes highly distinctive, robust class separation (severe morphological deviation).

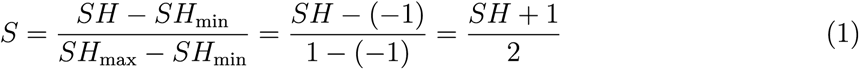

For the representative Test Subset 1, the derived Severity Factor *(S)* was approximately 0.66 for the intermediate U-EC-09 class and 0.76 for the highest U-EC-11 class, computed relative to the uninoculated baseline (U-EC-00). Notably, across all independent test subsets, *S* consistently registered within a bounded range of [0.6, 0.8]. This trend mathematically demonstrates that higher *E. coli* concentrations yield a greater severity factor, reflecting the severe morphological pattern deviation caused by increased bacterial presence. Consequently, these results validate that our derived severity factor can potentially translate the pattern deviation caused by bacterial load into a robust computational metric.

Although a comprehensive evaluation across a broader spectrum of bacterial concentrations is necessary for rigorous validation, the current study firmly establishes this severity factor for examining pattern deviation using feature cluster analysis. It is important to note that, at this foundational stage, *S* is formulated as a qualitative metric rather than interpreting the absolute magnitudes. Nevertheless, we introduce this metric to demonstrate that image-based pattern analysis can be used for understanding diagnostic severity, suggesting a valuable tool for diagnostic support.

## 4. Discussion

### 4.1. Feature Cluster Analysis

Both global performance metrics and low-dimensional maps reveal feature cluster overlap at lower concentrations (U-EC-00 and U-EC-09). In this subsection, we investigate this overlap and the morphological distinctiveness across the three bacterial concentrations in further detail. For illustration, we evaluate two specific test subsets, whose representative patterns are shown in Fig. 7. The 2D t-SNE projections are presented in Fig. 8, with panels (A) and (B) corresponding to Test Subsets 1 and 2, respectively. Based on these projections, the extracted standardized features clearly separate the samples of the highest concentration (U-EC-11) from the lower concentrations. Furthermore, a distinct variation is visible between these subsets at lower concentrations. Specifically, Test Subset 2 exhibits significant feature overlap on these low-dimensional maps. While t-SNE is visualized here, this behavior is consistently observed using PCA as well.

**Figure 8:**
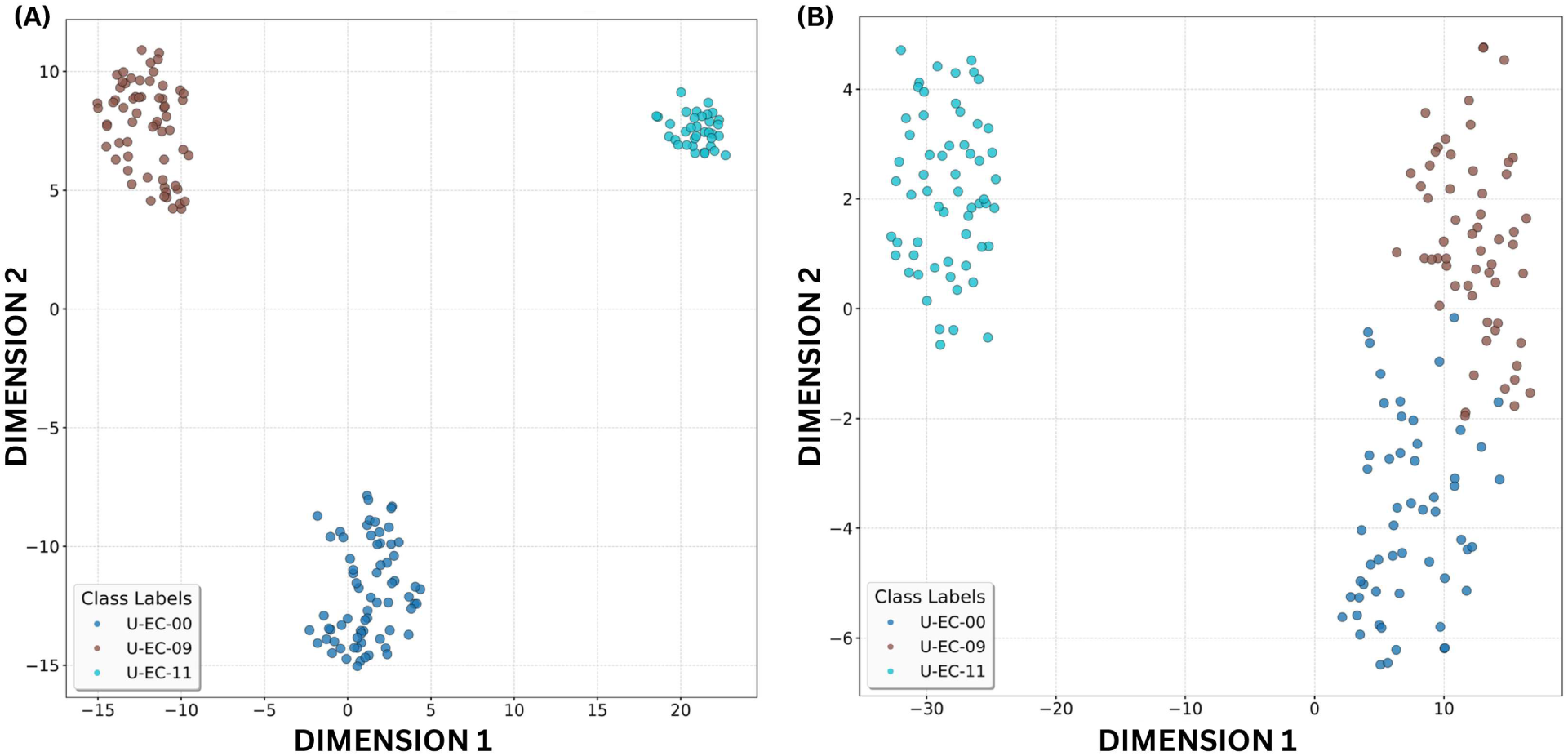
2D t-SNE plots for the two selected test subsets (Test Subset 1 and Test Subset 2), with one demonstrating clear class separation and the other showing partial overlap at lower concentrations, respectively.

The feature overlap observed between the uninoculated baseline (U-EC-00) and the 10^9^ CFU/mL concentration (U-EC-09) directly accounts for the comparatively lower classification performance. Furthermore, this overlap significantly impacts the CVIs utilized for computing the severity factor. Comparing Table 4 and Table 5, reveal an increase in the Davies-Bouldin index, alongside a decrease in the Silhouette score, Calinski-Harabasz index, and Centroidal Distance in the pair-wise metrics corresponding to U-EC-00 and U-EC-09. These metrics confirm the feature cluster overlap, effectively establishing the lower limit of morphological pattern distinctiveness for the current framework. This is further validated by additional dedicated experiments performed at even lower *E. coli* concentrations ^2^ (10^7^ and 10^8^ CFU/mL). However, crucially, the Severity Factor (S) still qualitatively reflects a consistent increase in morphological pattern deviation relative to the baseline as bacterial concentration increases.

**Table 4:** Comparison of Cluster Validity Indices (CVIs) for the U-EC-09 and U-EC-11 feature clusters against the U-EC-00 baseline, using standardized MobileNetV2 embeddings from Test Subset 1.

| CVI | U-EC-00 vs U-EC-09 | U-EC-00 vs U-EC-11 |
| --- | --- | --- |
| Silhouette Score | 0.343 | 0.506 |
| Normalized Silhouette Score | 0.671 | 0.753 |
| Davies-Bouldin Index | 1.236 | 0.759 |
| Calinski-Harabasz Index | 81.627 | 133.620 |
| Centroidal Distance | 32.227 | 45.450 |

**Table 5:** Comparison of Cluster Validity Indices (CVIs) for the U-EC-09 and U-EC-11 feature clusters against the U-EC-00 baseline, using standardized MobileNetV2 embeddings from Test Subset 2.

| CVI | U-EC-00 vs U-EC-09 | U-EC-00 vs U-EC-11 |
| --- | --- | --- |
| Silhouette Score | 0.142 | 0.467 |
| Normalized Silhouette Score | 0.571 | 0.734 |
| Davies-Bouldin Index | 2.265 | 0.893 |
| Calinski-Harabasz Index | 20.833 | 135.602 |
| Centroidal Distance | 14.888 | 36.807 |

Incorporating multiple images within each subset ensured that the observed morphological variations reflect consistent, generalizable pattern formations, rather than localized noise or experimental artifacts due to single datapoint predictions at a given bacterial concentration. While the current study establishes a robust methodological foundation, future investigations will focus on scaling the dataset and varying substrate topographies and ambient conditions to further elucidate these pattern formation dynamics. Crucially, this subset-centric approach motivated our shift from discrete, single datapoint predictions to a continuous, cluster-level analysis within the feature space for severity estimation. As this framework transitions toward clinical application with real-world patient data encompassing multiple complications, this cluster-based approach will be pivotal. By evaluating morphological pattern overlap and distinctiveness within the feature space, complex morphological behaviors can be correlated across diverse clinical scenarios. This mathematically grounded severity estimation procedure exhibits strong potential for direct extension to diverse bacterial pathogens and alternative complex biofluids, establishing a highly adaptable, real-time diagnostic tool at the point of care.

### 4.2 Ablation Study

Given our limited dataset size, lighter convolutional backbones (models with fewer parameters) were prioritized for feature extraction to avoid overfitting. The primary deep convolutional backbone was selected from multiple architectures across various CNN families through a systematic ablation study, including ResNet-34, ResNet-50 [8], Xception [72], EfficientNet-BO [9], RepVGG-AO [73], DenseNet-121 [10], MobileNetV2 [11], and MobileOne-SO [74]. All models were assessed using a linear classifier head (described in Sec. 2.3) to determine their predictive capabilities. The macro-averaged global performance metrics for these architectures are presented in Table 6. Notably, MobileNetV2 achieved a peak accuracy of 92.5%, significantly outperforming heavy-weight architectures like Xception and ResNet-50 on our dataset. Consequently, MobileNetV2 was established as the primary backbone for this study. Crucially, its low memory footprint and minimal computational overhead make it exceptionally well-suited for eventual deployment in resource-constrained, embedded Point-of-Care diagnostic systems. Subsequently, a suite of secondary classifiers was evaluated for robustness analysis, solely within this chosen model backbone. Importantly, this feature extraction backbone can be seamlessly replaced with any other architecture, highlighting the architectural versatility of the framework for diverse scenarios.

**Table 6:**
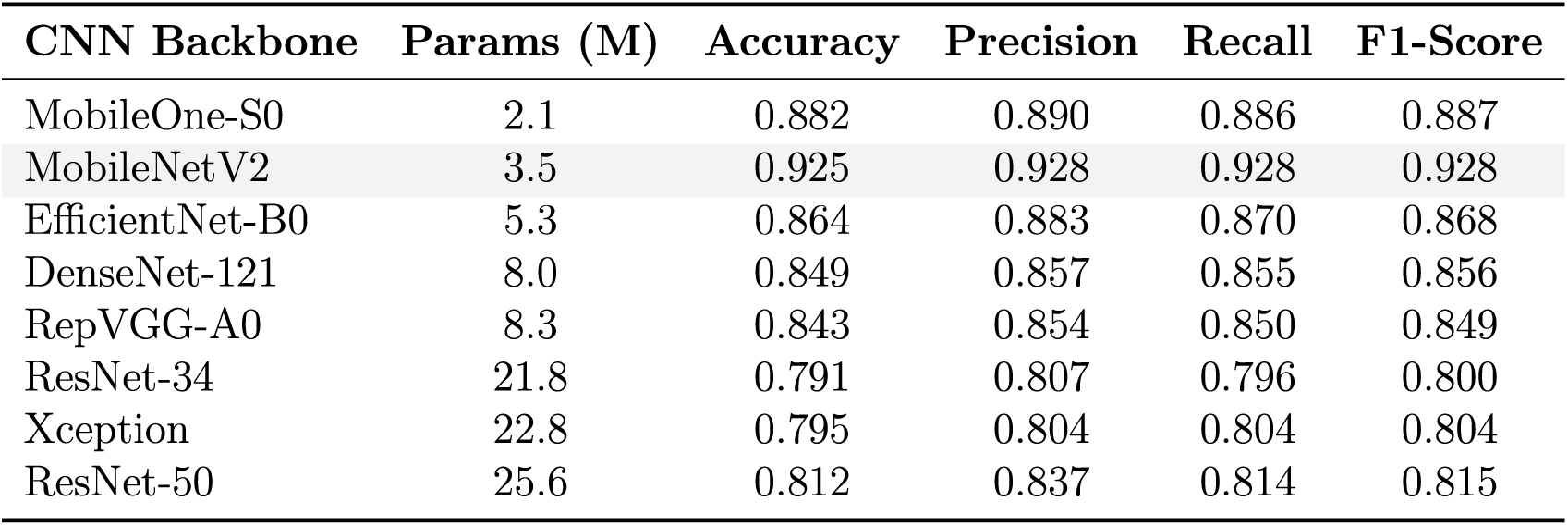
Comparison of model parameters and macro-averaged performance metrics across multiple deep convolutional architectures using a standardized linear classification head on the held-out test fold.

| CNN Backbone | Params (M) | Accuracy | Precision | Recall | F1-Score |
| --- | --- | --- | --- | --- | --- |
| MobileOne-S0 | 2.1 | 0.882 | 0.890 | 0.886 | 0.887 |
| MobileNet V2 | 3.5 | 0.925 | 0.928 | 0.928 | 0.928 |
| EfficientNet-B0 | 5.3 | 0.864 | 0.883 | 0.870 | 0.868 |
| DenseNet-121 | 8.0 | 0.849 | 0.857 | 0.855 | 0.856 |
| RepVGG-A0 | 8.3 | 0.843 | 0.854 | 0.850 | 0.849 |
| ResNet-34 | 21.8 | 0.791 | 0.807 | 0.796 | 0.800 |
| Xception | 22.8 | 0.795 | 0.804 | 0.804 | 0.804 |
| ResNet-50 | 25.6 | 0.812 | 0.837 | 0.814 | 0.815 |

## 5. Conclusion

In this study, we introduced a machine learning-based framework for morphological pattern analysis and severity estimation using dried deposits formed by *E.* coZz-laden sessile urine droplets inoculated at three distinct bacterial concentrations. Utilizing a lightweight deep convolutional backbone as a spatial feature extractor, we first performed ternary classification based on the bacterial concentration. This classification performance was visually interpreted by projecting the high-dimensional feature embeddings onto lower-dimensional maps using linear and non-linear dimensionality reduction techniques. Furthermore, we formulated a novel qualitative severity factor by applying CVIs to these standardized feature vectors. This approach evaluates the morphological pattern deviation caused by bacterial presence as a subset-specific measure. Critically, because the computational inference time of the trained model is negligible, the total diagnostic turnaround is limited solely by the optical imaging and evaporation processes, establishing its viability for rapid deployment. To enhance predictive generalization and for robust diagnosis, future investigations will focus on incorporating the current framework across a broader spectrum of bacterial concentrations and real-world uropathogenic patient samples. Integrating this framework into mobile or cyber-physical embedded platforms will transform its predictive capabilities into a practical, real-time diagnostic tool, delivering an accessible and cost-effective solution for resource-limited clinical environments.

## Data Availability

All data produced in the present study are available upon reasonable request to the authors.

## 6. Acknowledgments

A.G. acknowledges SERC (Supercomputer Education and Research Center) for GPU support. S.B. acknowledges support from the INAE (Indian National Academy of Engineering) Chair Professorship and J. C. Bose Grant of the Anusandhan National Research Foundation (ANRF). We would like to express our sincere thanks to Mr. Anmol Singh, Mr. Mradul Namdeo, and Mr. Raghavendra K N from the Indian Institute of Science for their contribution to the experimental studies.

## 7. CRediT authorship contribution statement

- Saptarshi Basu (S.B.): Conceptualization, Writing – review & editing, Resources, Funding acquisition.
- Dipshikha Chakravortty (D.C.): Conceptualization, Writing - review & editing.
- M Ashwin Ganesh (A.G.): Conceptualization, Project administration, Methodology, Formal analysis, Investigation, Software. Visualization, Validation, Supervision, Writing - original draft, Writing - review & editing.
- Sophia M (S.M.), Jason Joy Poopady (J.J.), Abdur Rasheed (A.R.), Durbar Roy (D.R.), Arney Nitin Agharkar (A.A), Kirti Parmar (K.P): Data curation, Methodology, Investigation.
- Visakh Vaikuntanathan (V.V.): Formal analysis.

## Footnotes

1 A preliminary geometry-based feature analysis performed to study the variation in pattern formation behavior has been provided in Section 1 of the supplementary material.

2 Detailed results from pattern classification and low-dimensional maps performed at *E. coli* concentrations of 10^7^ and 10^8^ CFU/mL are provided in Section 2 of the supplementary material.

