## Supplementary Material for "Deep Learning-Driven Pattern Analysis of Dried *E. coli*-Laden Urine Deposits for Point-of-Care Diagnostics"

### Supplementary Material: Deep Learning-Driven Pattern Analysis of Dried Deposits formed by *E. coli*-Laden Urine Droplets for Point-of-Care Diagnostics

M Ashwin Ganesh, Sophia M, Jason Joy Poopady, Abdur Rasheed, Durbar Roy, Amey Nitin Agharkar, Visakh Vaikuntanathan, Kirti Parmar, Dipshikha Chakravortty, Saptarshi Basu

#### 1 Geometry-Based Features: As a Starting point

Based on preliminary visual observations, the dried deposits formed by *E. coli*-laden sessile urine droplets seemed to show variability in the underlying pattern formation at various inoculated concentrations. As a first step to decode this pattern formation behavior, a geometry-based approach was performed with a pertinent question in mind: Could we identify and extract features that contain the morphological signature of the specific bacterial concentration? Therefore, as a preliminary analysis, a representative set of experimental microscopic images was considered, and certain geometry-based features were evaluated. A few of the estimated features studied include: number of particles ($N$); areal number density of particles within the deposit footprint area ($N/A_0$); total area of particles ($A_{tot}$) normalized by the area of the deposit footprint ($A^* = A_{tot}/A_0$); total area of particles normalized by the maximum possible area within the image ($A^{**} = A_{tot}/A_{im}$); total perimeter of particles ($P_{tot}$) normalized by the perimeter of the deposit footprint ($P^* = P_{tot}/P_0$); total perimeter of particles normalized by the maximum possible perimeter within the image ($P^{**} = P_{tot}/P_{im}$); and shape factor ($\Psi = A_{tot}/P_{tot}^2$).

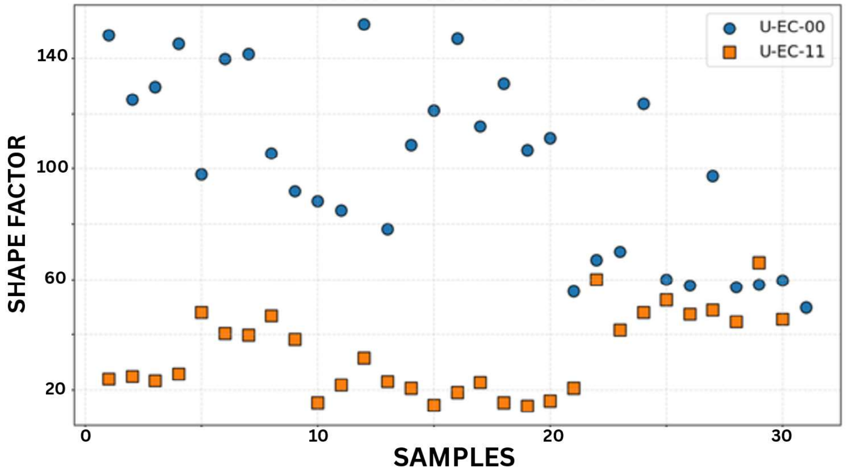


Figure 1: Variation of Shape factor $\Psi = A_{tot}/P_{tot}^2$ for representative samples with presence of E.coli.

Based on this evaluation, most of the geometry-based features showed a distinction between the extreme scenarios: the uninoculated baseline U-EC-00 and the highest concentration U-EC-11. The variation in the shape factor corresponding to these scenarios has been depicted in Fig. 1. Though the subtle variations across all intermediate concentrations were not completely evident through these preliminary features, there remained a tangible possibility for understanding the patterns with a comprehensive analysis. Motivated by this preliminary study, the necessary next step was to step into unveiling the variation in pattern formation by employing deep learning-based methodologies, utilizing their capacity to identify intrinsic details to uncover patterns that are not trivially perceptible to humans.

#### 2 Morphological Pattern Analysis at Lower Concentrations

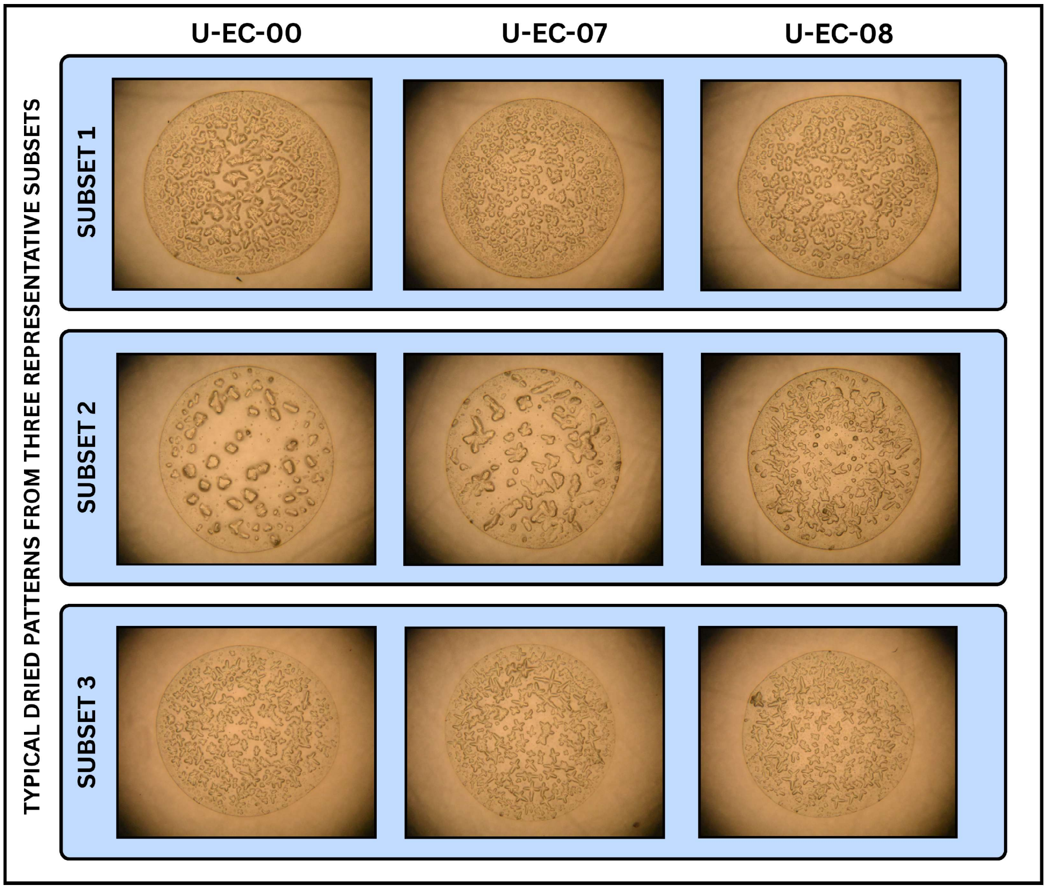


Figure 2: Representative images of the typical final dried deposit patterns at lower *E. coli* concentrations.

In addition to the pattern analysis at higher concentrations described in the main text, we performed ternary pattern classification and severity estimation at significantly lower bacterial concentrations to establish the physical Limit of Detection (LoD) of the framework. Analogously, the microscopic images were organized into experimental subsets, wherein each subset comprised three distinct bacterial concentration classes: U-EC-00, U-EC-07, and U-EC-08, averaging 60 images per class. This yielded a total of 3,745 images, distributed as 1,225 for U-EC-00, 1,236 for U-EC-07, and 1,284 for U-EC-08, all stored in high-resolution (6000 × 4000 pixels) RGB format. Importantly, the urine samples comprising this dataset are completely independent of those utilized in the high-concentration pattern analysis. Representative deposits at each bacterial concentration from three selected subsets are depicted in Fig. 2. To prevent data leakage, the data were partitioned at the subset level: 17 subsets were allocated for training and validation, while the remaining 4 were strictly reserved as a held-out test set.

Utilizing MobileNetV2 as the primary feature extractor, the ternary classification of morphological patterns was evaluated on the held-out test set (Fig. 3A). For the deposits at these lower *E. coli* concentrations, the overall accuracy expectedly dropped to approximately 53%, with macro-averaged precision, recall, and F1-scores ranging between 53% and 58%.

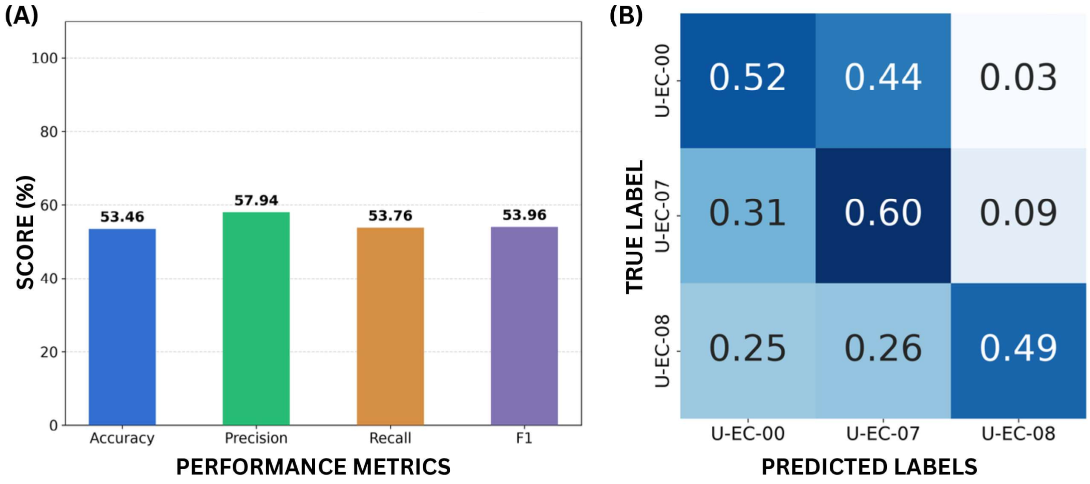


Figure 3: (A) The macro-averaged performance metrics and (B) the row-normalized confusion matrix for morphological pattern classification, based on the linear head classifier, evaluated at lower concentrations.

The class-wise evaluation (Table 1) and the row-normalized confusion matrix (Fig. 3B) demonstrate this drastic drop in predictive ability. This pattern behavior is visually corroborated by the 2D PCA and t-SNE mappings (Fig. 4). Specifically, the t-SNE maps indicate severe, subset-level topological overlap for these lower-concentration deposits.

Table 1: Class-wise performance metrics utilizing the linear head classifier.

| Class | Precision | Recall | F1-Score |
|---|---|---|---|
| U-EC-00 | 0.486 | 0.523 | 0.504 |
| U-EC-07 | 0.434 | 0.604 | 0.505 |
| U-EC-08 | 0.818 | 0.486 | 0.610 |

To validate the classifier-agnostic robustness of the extracted feature embeddings at these lower concentrations, the standardized feature vectors were independently evaluated across a suite of secondary machine learning algorithms (Table 2). Predictive performance performance remained in a consistently lower range across all chosen classifiers, with Logistic Regression achieving a maximum overall accuracy of about 59.3%. This suggests that the reduction in discriminative ability reflects the lack of distinct morphological features rather than an artifact of algorithmic choice.

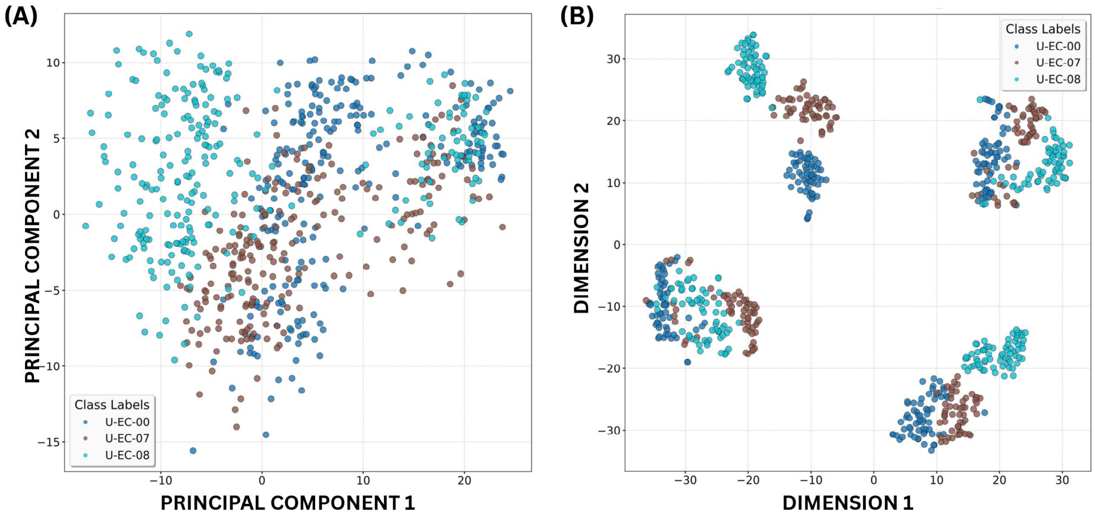


Figure 4: Two-dimensional (A) Principal Component Analysis (PCA) and (B) t-SNE maps based on the standardized feature vectors.

Table 2: Comparison of macro-averaged performance metrics evaluated across multiple classifiers using the standardized feature embeddings obtained from the MobileNetV2 model on the held-out test fold.

| Classifier | Accuracy | Precision | Recall | F1-Score |
|---|---|---|---|---|
| Support Vector Machine | 0.555 | 0.584 | 0.557 | 0.560 |
| Logistic Regression | 0.593 | 0.630 | 0.590 | 0.592 |
| K-Nearest Neighbors | 0.550 | 0.583 | 0.551 | 0.554 |
| Random Forest | 0.543 | 0.589 | 0.546 | 0.548 |
| Naive Bayes | 0.551 | 0.607 | 0.555 | 0.558 |

Furthermore, severity estimation was evaluated using a representative test subset (Fig. 5). Crucially, although there is heavy global feature overlap, the cluster validity indices (Table 3) still quantitatively reflect an increase in pattern deviation relative to the uninoculated baseline as bacterial concentration increases. This confirms that the Severity Factor remains qualitatively indicative of pattern deviation, despite the reduced predictive ability observed in pattern classification.

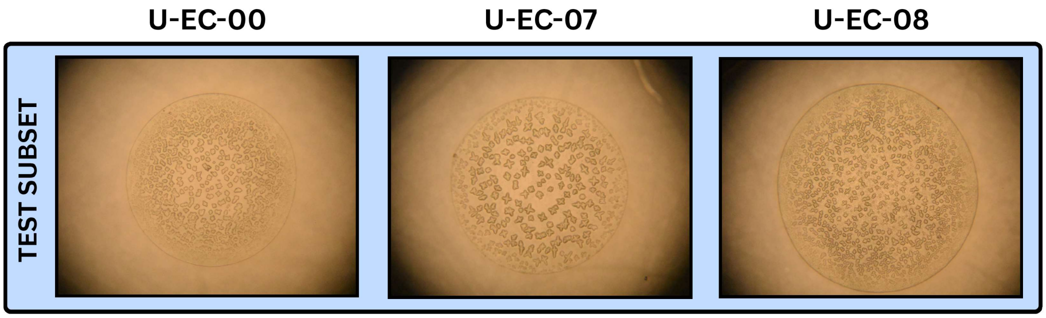


Figure 5: Representative patterns from the selected test subset.

Table 3: Comparison of Cluster Validity Indices (CVIs) for the U-EC-07 and U-EC-08 feature clusters relative to the baseline U-EC-00, evaluated based on the extracted MobileNetV2 feature embeddings following standardization, for the selected test subset.

| **CVI** | **U-EC-00 vs U-EC-07** | **U-EC-00 vs U-EC-08** |
|---|---|---|
| Silhouette Score | 0.116 | 0.205 |
| Normalized Silhouette Score | 0.558 | 0.602 |
| Davies-Bouldin Index | 2.632 | 1.827 |
| Calinski-Harabasz Index | 15.421 | 35.546 |
| Centroidal Distance | 13.428 | 18.378 |

The sharp drop in predictive ability observed below U-EC-09 globally establishes the Limit of Detection (LoD) for our framework. To validate this boundary further, future investigations will focus on experiments with a more diverse spectrum of sample collections and bacterial concentrations.
